# The Presence of Non–organ-specific Autoantibodies in Chronic Hepatitis C Treatment-naïve Patients

**DOI:** 10.1101/2022.01.07.22268924

**Authors:** Luis Jesuíno de Oliveira Andrade, Ingrid Silva Santos Padilha, Luis Matos de Oliveira, Gabriela Correia Matos de Oliveira, Raymundo Paraná

## Abstract

**Background:** In the patients with hepatitis C virus (HCV) various immune-mediated phenomena are described, and non–organ-specific autoantibodies (NOSAs) in particular are common. The aim of the present study was to investigate the NOSAs prevalence in chronic hepatitis c treatment-naïve patients.

**Patients and Methods:** Sera of 76 consecutive HCV treatment-naïve patients were considered to be eligible for this study for evaluation of Anti-nuclear, anti-smooth muscle, anti-mitochondrial, anti-neutrophil-cytoplasmatic and anti-liver/kidney microsomal antibodies. Criteria of eligibility were serum anti-HCV antibody and HCV RNA positivity, chronic inflammation revealed by histological analysis of the liver, genotyping, treatment-naïve patients, and no have the diagnosis of probable or definite autoimmune hepatitis.

**Results:** Mean chronological age for the 76 patients (44 females and 32 males) was 51.3 ± 13,9 years (range: 20-67 years). Nineteen patients (25.0%) infected with HCV had detectable levels of NOSAs at before combined antiviral treatment. SMA was present in 16 (21.0%) of 76 patients, ANA in 2 patients (2.6%), and pANCA (perinuclear ANCA) in 1 patients (1.3%). No patient had specimens reactive to AMA, LMK, or cANCA (cytoplasmic ANCA).

**Conclusions:** In this study, we show the NOSAs positivity in chronic hepatitis c treatment-naïve patients. Assigned to high prevalence of SMA positivity is associated with high METAVIR score, and HCV-genotype 1 and 1b, may reflect a release of intracellular antigens at the time of hepatocellular injury triggering immune responses in the form of autoantibody production or a direct infection of immunocytes by the HCV.

## Introduction

In the patients chronically infected by hepatitis C virus (HCV) various immune-mediated phenomena have been frequently described, and non–organ-specific autoantibodies (NOSAs) in particular are common examples of autoreactivity in a considerable number of individuals with acute and chronic hepatitis C.^1-3^ The NOSAs includes anti-nuclear antibodies (ANA), anti-smooth muscle antibodies (SMA), anti-mitochondrial antibodies (AMA), anti-neutophil-cytoplasmatic antibodies (ANCA), and anti-liver/kidney microsomal antibodies (LKM).

The frequency of NOSAs in HCV-related chronic hepatitis ranges from 10% to 21% for ANA, 10% to 55% for SMA, 0 to 8% for AMA, 0 to 39% for ANCA and 0% to 6% for LKM1,^1^ and is associated with both cirrhosis and older age.^4^ The high prevalence of NOSAs may reflect a chronic antigenic stimulation or a direct infection of immunocytes by the HCV.^5,6^

The aim of the present study was to investigate the NOSAs in patients chronically infected by HCV before treatment with interferon-alpha and ribavirin.

## Patients and Methods

### Study patients

Sera of 76 consecutive HCV treatment-naïve patients were considered to be eligible for this study. Criteria of eligibility were serum anti-HCV antibody and HCV RNA positivity, chronic inflammation revealed by histological analysis of the liver, genotyping, treatment-naïve patients, and no have the diagnosis of probable or definite autoimmune hepatitis.

All the patients gave their informed consent to participate to the study, and the study was approved by the ethical committee of State University of Santa Cruz, Bahia, Brazil in accordance to the Declaration of Helsinki.

### Virological assays

The detection of antibodies to HCV (Anti-HCV) was measured by means of an enzyme-linked immunosorbent assay, using structural and non-structural HCV antigens (AXSYM System; Abbott Laboratories, Chicago, IL, USA). HCV-RNA load were available by quantitative polymerase chain reaction (PCR) using primers derived from the highly conserved 5’ noncoding region of the viral genome (Amplicor® HCV Detection KIT V2.0; Roche Molecular Systems Inc., Somerville, NJ, USA). HCV-genotypes were performed by PCR amplification of the core region of the HCV genome with specific antisense primers (Applied Biosystems, Foster City, CA). The classification of the genotypes was carried out according to Simmonds et al.^7^

### Liver Histology

All subjects who were found positive for HCV-RNA underwent percutaneous liver biopsy and histological parameters were classified according to the score system of inflammation activity and fibrosis.

The degree of histological fibrosis was using the METAVIR score. The fibrosis is graded on a 5-point scale from 0 to 4. The activity, which is the amount of inflammation (specifically, the intensity of necro-inflammatory lesions), is graded on a 4-point scale from A0 to A3. Fibrosis score F0= absence of fibrosis; F1= fibrous expansion of portal areas; F2= portal to portal bridging fibrous tracts; F3= portal-central bridging fibrous septa; F4= cirrhosis (bridging fibrous septa with parenchymal nodules). Activity score: 0 = no activity; A1 = mild activity; A2 = moderate activity; A3 = severe activity.^8,9^

### NOSAs determination

The presence of ANA, SMA, AMA and LKM1 were detected by indirect immunofluorescence (IIF) at serum dilutions with positive titre ≥ 1:40. Tests for detection of ANCA were detected by IIF on ethanol-fixed granulocytes with positive titre ≥ 1:20. The specificity, reproducibility and optimal conditions of these assays have been determined in preliminary experiments. Autoantibody titers were quantified with commercially available kits for IIF (INOVA, San Diego, CA).

### Statistical analysis

The comparison of the association between categorical groups was performed using chi-square or Fischer’s exact test. The nonparametric Mann–Whitney test was used for comparison the difference between groups. A p-value < 0.05 was considered significant.

## Results

Mean chronological age for the 76 patients (44 females and 32 males) was 51.3 ± 13,9 years (range: 20-67 years).

Liver biopsies were performed in all patient, the results on inflammatory activity and fibrosis (METAVIR score) are shown in Table 1.

**Table 1.** Histopathological features of patients positive for HCV-RNA

| Liver biopsy |  | Patients (n) |
| --- | --- | --- |
| Inflammatory activity |  |  |
| 0 |  | 22 |
| 1 |  | 20 |
| 2 |  | 29 |
| 3 |  | 5 |
| Fibrosis |  |  |
| F0 |  | 2 |
| F1 |  | 16 |
| F2 |  | 27 |
| F3 |  | 24 |
| F4 |  | 7 |

Nineteen patients (25.0%) infected with HCV had detectable levels of NOSAs at before combined antiviral treatment. SMA was present in 16 (21.0%) of 76 patients, ANA in 2 patients (2.6%), and pANCA (perinuclear ANCA) in 1 patients (1.3%), and and is reported in Table 2. No patient had specimens reactive to AMA, LMK, or cANCA (cytoplasmic ANCA).

**Table 2.**
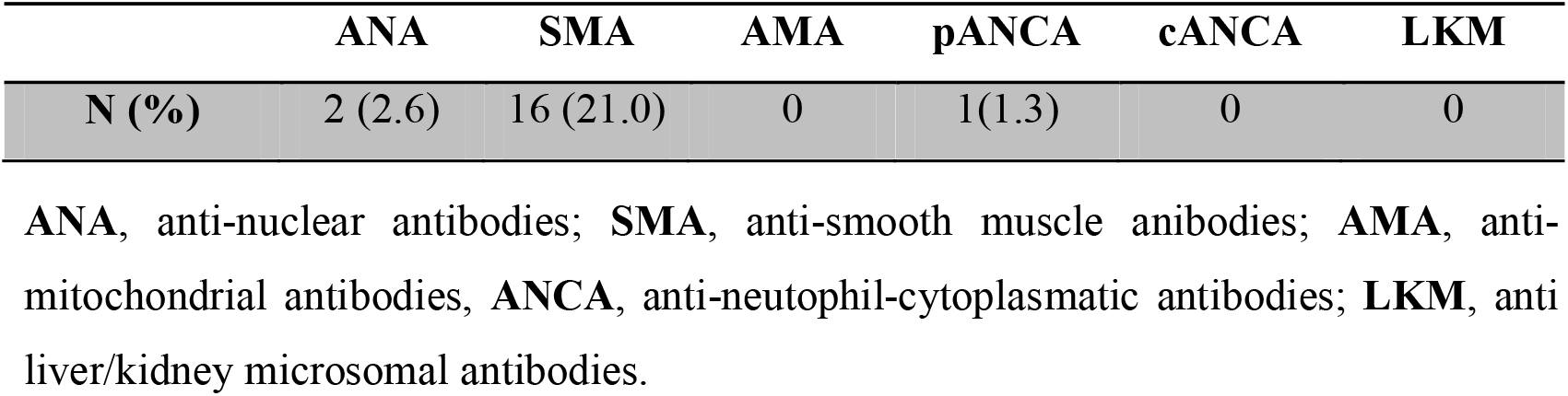
Prevalence of NOSAa in anti-HCV positive patients (n = 76).

The relationship between HCV genotypes and presence of NOSAs demonstrates that HCV-genotype 1 and 1b was significantly more frequent in NOSAs positive subjects, and is reported in Table 3.

**Table 3.**
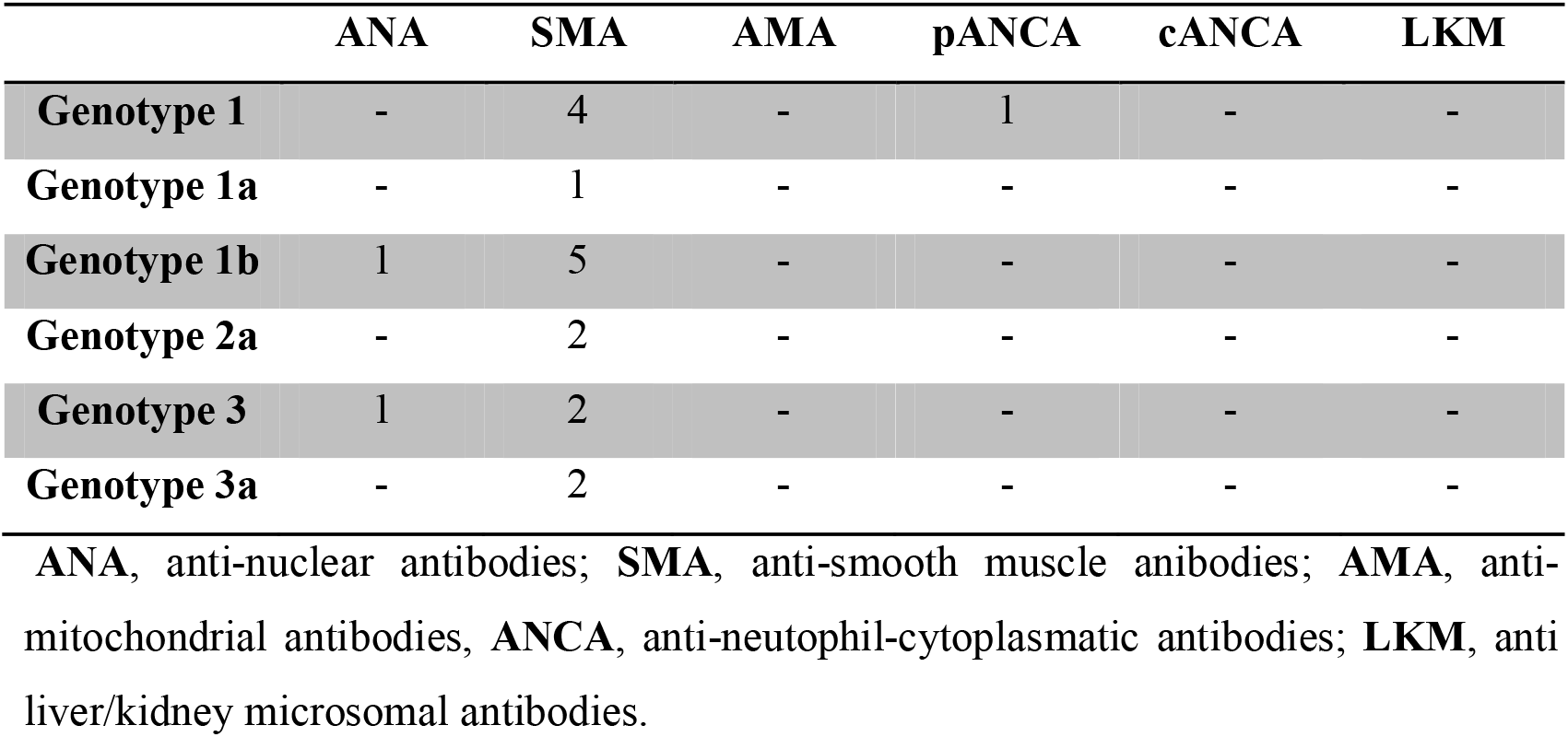
NOSAs and HCV genotype

|  | ANA | SMA | AMA | pANCA | cANCA | LKM |
| --- | --- | --- | --- | --- | --- | --- |
| **Genotype 1** | - | 4 | - | 1 | - | - |
| **Genotype 1a** | - | 1 | - | - | - | - |
| **Genotype 1b** | 1 | 5 | - | - | - | - |
| **Genotype 2a** | - | 2 | - | - | - | - |
| **Genotype 3** | 1 | 2 | - | - | - | - |
| **Genotype 3a** | - | 2 | - | - | - | - |

ANA, anti-nuclear antibodies; SMA, anti-smooth muscle antibodies; AMA, anti-mitochondrial antibodies, ANCA, anti-neutrophil-cytoplasmic antibodies; LKM, anti liver/kidney microsomal antibodies.

The correlation between NOSAs positivity and degree of inflammatory activity and the degree of fibrosis upon histological examination of the liver biopsies reveal a highly significant statistic (p= 0,002, and p= 0,001), while no significant correlation has been found between NOSAs positivity and age, sex.

## Discussion

NOSA have been frequently detected in sera of HCV-infected patients as reported by several authors.^1,10,11^ In this study, we demonstrated that the prevalence of NOSAs in chronic hepatitis C treatment-naïve patients was of 25.0%. Our study confirmed that NOSAs is frequently detected in HCV patients. Similarly to previous reports by Chrétien and colleagues,^12^ these autoantibodies appeared to be a part of the natural course of chronic hepatitis C. However, in previous reports from Europe and United States the prevalence of NOSAs is only slightly top than that observed in our study.^13,14^

In our study, the subjects had SMA with highest prevalence rates (21.0%), while ANA and pANCA showed low prevalence rates (2.6% and 1.3%, respectively). The prevalence of SMA in our study is comparable to previous studies, which demonstrated these antibodies to be present at similar frequencies.^3,15,16^ In contrast with some previous reports LKM were not detected in our report, most likely due to the sample size.^17,18^ AMA and cANCA, as shown in our study is comparable to previous reports have been rarely found in patients chronically infected by hepatitis C virus.^19,20^

The relationship between HCV genotypes and presence of NOSAs in this study demonstrates that HCV-genotype 1 and 1b was significantly more frequent in NOSAs positive subjects. Lenzi and colleagues,^15^ demonstrated a link between the presence of reactivity for SMA and HCV genotype 1b.

The association between patients chronically infected by HCV and the presence of NOSAs is still controversial. Cassani and colleagues,^21^ found an association between NOSAs (mostly ANA and SMA) and necro-inflammatory activity in liver biopsies. These results are strengthened by the observations obtained in this study, that show the correlation between NOSAs positivity and degree of inflammatory activity and the degree of fibrosis upon histological examination of the liver biopsies showing a highly significant statistic. The study of Cassani and colleagues,^21^ also found an association between NOSAs and female gender, in our study no significant correlation has been found between NOSAs positivity and age, sex.

Therefore, previous studies^15,21^ along with our findings, strengthens the argument that continuous liver damage and hepatocyte necrosis in patients with chronic hepatitis C favors autoimmune abnormalities, and NOSAs also play an relevant role in the progression of stratification of liver damage, but urgent need of sufficient data with several prospective studies in order to definitely clarify to provide an answer.

## Conclusions

In this study, we show the NOSAs positivity in chronic hepatitis c treatment-naïve patients. Assigned to high prevalence of SMA positivity is associated with high METAVIR score, and HCV-genotype 1 and 1b, may reflect a release of intracellular antigens at the time of hepatocellular injury triggering immune responses in the form of autoantibody production or a direct infection of immunocytes by the HCV.

## Data Availability

All data produced in the present study are available upon reasonable request to the authors

